# Analysis of Blood Gene Expression Data Toward Early Detection of Alzheimer’s Disease

**DOI:** 10.1101/2021.07.26.21261147

**Authors:** Hamed Taheri Gorji, Ramtin Kardan, Neda Rezagholizadeh

## Abstract

Alzheimer’s Disease (AD) is a progressive neurodegenerative disorder and the most commonly diagnosed cause of dementia, and it is the fifth leading cause of death among people aged 65 and older. During the years, the early diagnosis of AD patients has been a significant concern for researchers, in view of the fact that early diagnosis not only can lead to saving lives of the AD patients but also could bring a considerable amount of saving in health and long-term care expenditures for both people and the government. Mild cognitive impairment (MCI), defined as a transitional state between being healthy and having AD, is considered an established risk factor for AD. Hence, an accurate and reliable diagnosis of MCI and, consequently, discrimination between healthy people, MCI individuals, and AD patients can play a crucial role in the early diagnosis of AD. In recent years, analysis of blood gene expression data has been grabbed more attention than the conventional AD diagnosis method because it provides the opportunity to investigate the biochemical pathways, cellular functions, and regulatory mechanisms for finding the key genes associated with MCI and AD. Therefore, in this study, we employed blood gene expression data from Alzheimer’s Disease Neuroimaging Initiative (ADNI), two feature selection methods for determining the most prominent genes related to MCI and AD, and three classifiers for the most accurate discrimination between three groups of healthy, MCI and AD. The proposed method yielded the selection of top ten genes from more than 49,000 genes and the best overall classification result between healthy and AD patients with average values of the area under the curve (AUC) of 0.77 ± 0.08. Furthermore, gene ontology (GO) analysis revealed that four genes were enriched with the GO terms of regulation of cell proliferation, negative regulation of cell population proliferation, signaling receptor binding, biological adhesion, and cytokine production.

## Introduction

Alzheimer’s disease (AD) is the most common diagnosed cause of dementia in older people [1]. Alzheimer’s is a progressive neurodegenerative disorder that causes memory loss and other adverse and potentially fatal symptoms. In recent years, several investigations have been conducted to predict this disease in patients at earlier stages. The early detection of AD can be beneficial by providing an opportunity for the patients to be considered for clinical trials. Furthermore, early detection can give patients enough time to plan for their financial and medical decisions.

Mild cognitive impairment (MCI) is a transitional stage between AD and a normal functional brain at a certain age [2]. and it is considered an early sign of Alzheimer’s disease. The accurate diagnosis of MCI condition helps the prognosis of AD more effectively.

The number of AD patients is exponentially increasing over the years, and based on estimation, around 13.8 million Americans will suffer from AD by 2050 [3]. With the rapid increase in the number of AD patients and the slow progress in treatment and prevention methods over the years, it is essential to investigate different approaches for finding any potential solution for more effective treatment or prevention for this disease.

Brain imaging modalities such as magnetic resonance imaging (MRI) and positron emission tomography (PET) have led to tremendous development in analyzing the brain and understanding brain function and its changes during the MCI and AD stages [4, 5]. Although using brain imaging techniques can provide vital information, these techniques are usually costly. Investigation to find an alternative approach for gathering the brain data from the patients to compare with normal people can be beneficial.

In recent years, human blood gene expression data have been received much attention as an affordable alternative approach and the appropriate method for monitoring and diagnosing AD and MCI [6-8].

One of the primary blood gene expression datasets available for researchers belongs to Alzheimer’s Disease Neuroimaging Initiative (ADNI). This dataset has been used in some recent studies. The authors of [9] conducted a study on ADNI and two other datasets, namely ANM1, and ANM2 to evaluate the gene expression relation between normal and the AD group. In this study, they selected high-quality RNA samples (RIN>=6.9) and then filtered out all the probes with a lower value than median gene expression to decrease the background noise. Besides, they normalized the datasets and selected the DEGs using significant analysis of microarrays (SAM). Eventually, they applied variational autoencoder (VAE), selected six genes to be trained in a machine learning model, and observed significant expression variations in genes enriched with inflammatory or immune pathways. However, predictive capacity has varied considerably from study to study[9, 10]. In another study based on the ADNI dataset [10]. the authors investigated the application of network medicine (NM) on AD. More specifically, the authors conducted a network-based analysis of the gene expression level and observed the co-expression patterns between the blood samples of NL, MCI, and AD.

Although gene expression data is a beneficial and affordable technique, its size is usually huge and makes the analysis too challenging. To reduce the amount of computation, and the size of data, selecting a suitable approach for eliminating the unrelated or redundant variables is vital. Hence, feature selection techniques can be very beneficial to understand the data and facilitate their visualization [11]. It is also helpful in decreasing the space for data storage and reducing computational expenses by decreasing training and utilization times. Since some techniques put more stress on one aspect of the selection; thus, it is crucial to choose the feature selection wisely [12].

The current study is directed toward research on human blood gene expression data from the ADNI database to determine the most relevant genes related to MCI and AD. Due to the massive volume of the ADNI data, we used two different feature selection techniques for selecting the most relevant genes related to MCI and AD, and also, we used three classifiers to discriminate between normal people, MCI individuals, and AD patients using the selected genes. Moreover, gene ontology was used to interpret the top selected genes’ function at the molecular and cellular levels.

## Material and Methods

The data used for this study belongs to Alzheimer’s Disease Neuroimaging Initiative (ADNI) database (adni.loni.usc.edu) [13]. The ADNI was launched in 2003 as a public-private initiative headed by Chief Investigator Michael W. Weiner, MD. The overall purpose of the ADNI was to provide a database composed of neuroimaging data such as MR and PET scan images, clinical data, genetic and biospecimen data that can be used by researchers for further understanding the mild cognitive impairment (MCI) and AD. The ADNI’s gene expression data extracted from blood samples of 811 participants from the ADNI WGS cohort using the Affymetrix Human Genome U219 Array (Affymetrix (www.affymetrix.com), Santa Clara, CA). Robust Multichip Average (RMA) normalization approach was used to pre-process the raw expression values derived directly from the CEL files.

Moreover, the blood RNA samples of 64 participants were removed from the dataset because they did not pass the quality control check. The final ADNI gene expression data was composed of 747 participants and 49,386 probe sets per participant mapped and annotated with reference to the human genome (hg19). All the participants were labeled with the diagnosis status of normal (NL), MCI, AD, NL to MCI, MCI to NL, and MCI to AD. In this study, we only considered the participants labeled with NL, MCI, and AD (stable status), and the participants who were diagnosed and labeled with the transitional states were removed. Finally, 713 participants (244 NL, 371 MCI, and 98 AD) were selected for further analysis. Subjects’ characteristics are shown in Table 1.

**Table 1.**
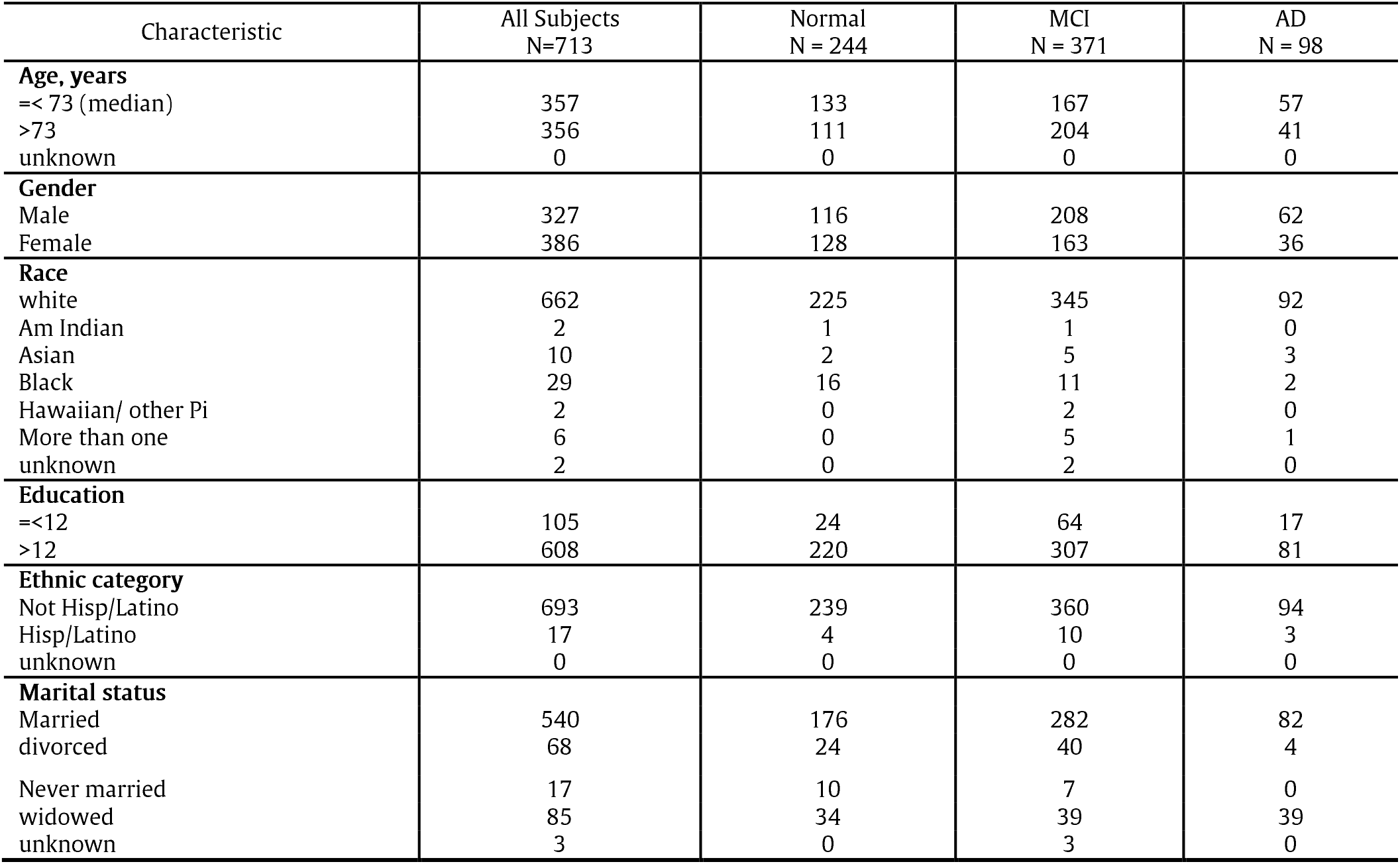
Subjects’ Characteristics in ADNI Blood Gene Expression Dataset

### 1) study Flowchart

To achieve the goals of the current work, we followed the chart illustrated in fig.3.

**Figure 1.**
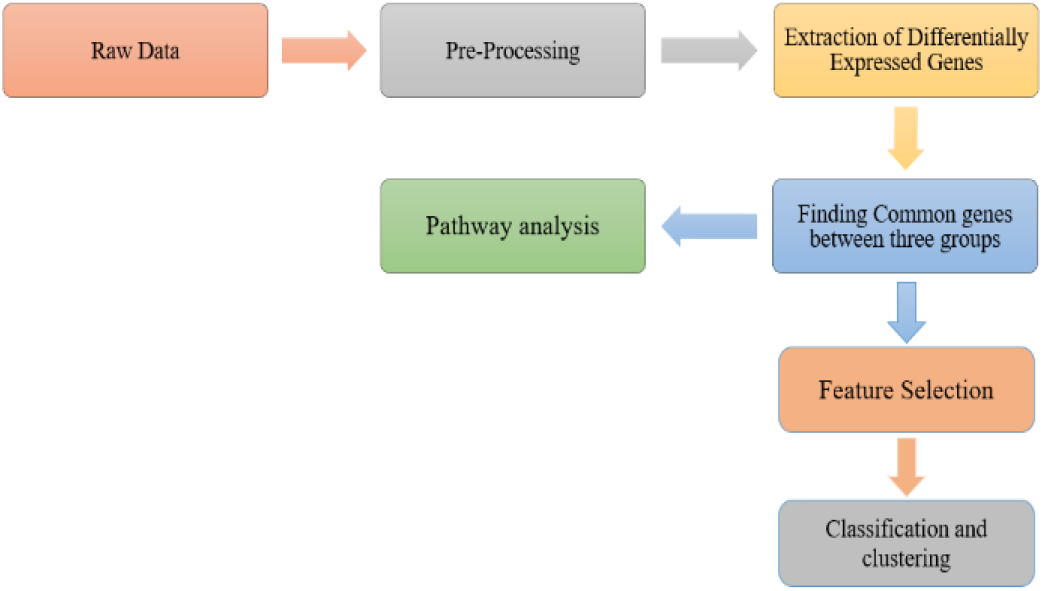
Prediction of Alzheimer’s disease using blood gene expression data flowchart

**Figure 2.**
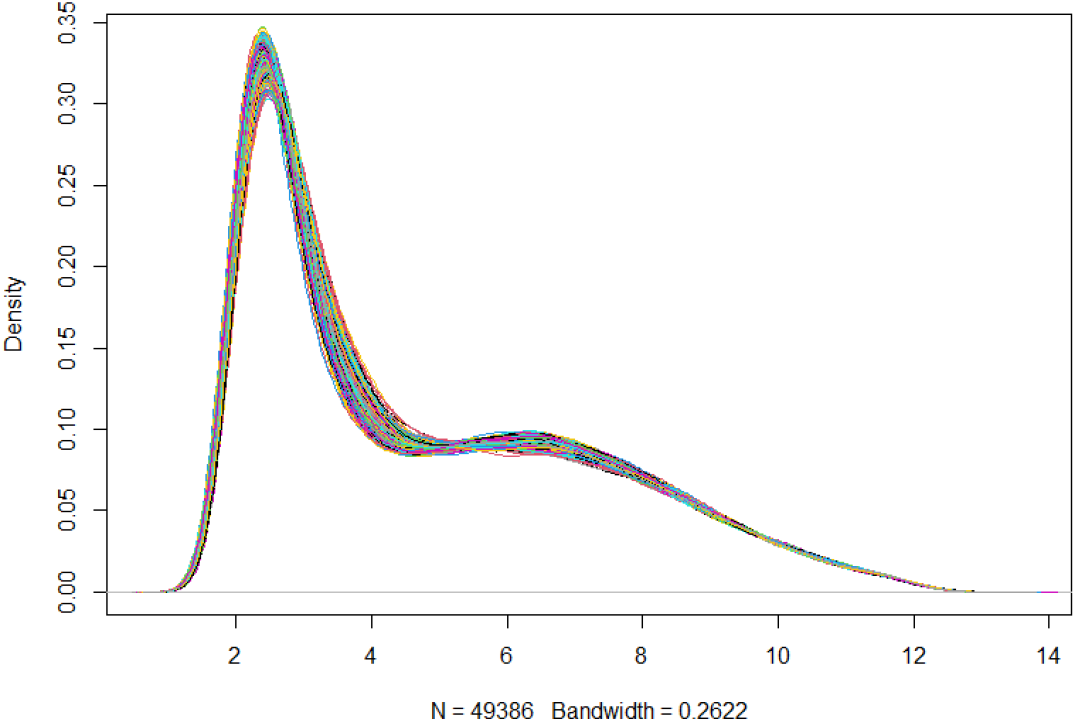
Kernel density estimation plot

**Figure 3:**
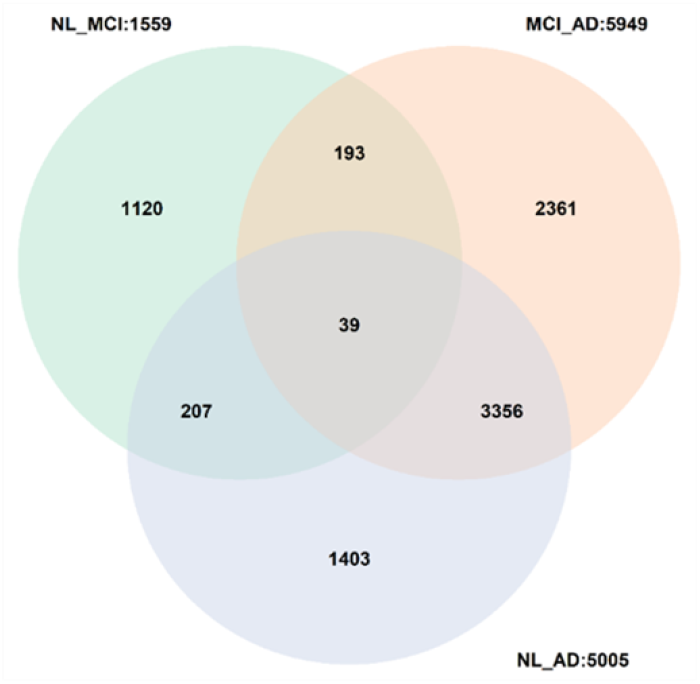
The Venn Diagram of the most expressed genes and 39 common genes.

### A. Pre-Processing

#### 1) Kernel Density estimation

Since the ADNI gene expression data were already normalized and log-scaled using the RMA normalization technique, further normalization was not needed. However, for the assurance about the data normalization, the kernel density estimation (KDE) approach was employed, and the plot can be seen in fig.4.

**Figure 4:**
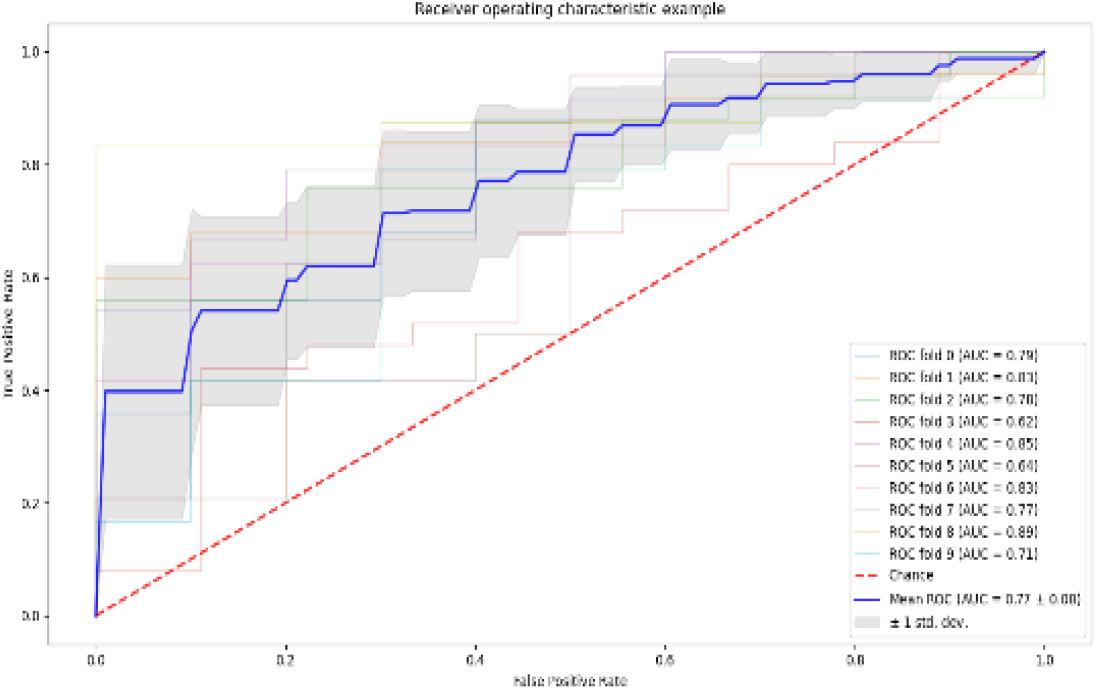
ROC curve for LassoCV and SVM

#### 2) Detection of differentially expressed genes

After evaluating the data normality, for detecting the differentially expressed (DE) genes between three groups of NL, MCI, and AD, Student’s t-test was used. For the t-test analysis, we utilized an unpaired t-test by considering the two-sided alternative hypothesis with a significance level of 0.05. The t-test was applied to the pairs of NL vs. MCI, NL vs. AD, and MCI vs. AD. After obtaining the p-value for each gene, all the p-values were adjusted using the false discovery rate (FDR) approach with the threshold level of 5 percent to limit the family error rate, and the genes with a p-value less than 0.05 were considered for further analysis.

#### 3) Feature Selection

Feature selection plays an essential role in data analysis. This method by reducing data dimensionality not only decreases the analyzing complexity and time but also can increase the performance of the model (accuracy improvement and reducing the overfitting) by removing the redundant and misleading features from data. Feature selection methods are often divided into three categories, namely filter-based, wrapper-based, and embedded methods. In this study, we used two feature selection methods: recursive feature elimination (RFE) and Lasso (Least Absolute Shrinkage and Selection Operator) cross-validation from wrapper-based and embedded methods, respectively [14, 15]. The RFE technique aims to find the best subset of performing features while generating models and storing the best or worst performing feature in a single iteration. The RFE can also make the next model by considering the left features before all features are exhausted. The Lasso feature selection method with the primary goal of reducing the prediction error is based on two main tasks, which are regulation and feature selection. By putting a penalty on the sum of the absolute value, this technique tries to minimize the sum of the square errors. The sum must be less than a specific threshold, and during the selection of the features, the variables with a non-zero coefficient may be selected as an integral part of the model after the shrinking process.

However, each feature selection algorithms ranks different features as the important features, and that is why we used machine learning classifiers to evaluate the efficiency of the selected features in differentiation between three groups of NL, MCI, and AD.

#### 4) Classification techniques

Classification as a supervised machine learning technique tries to learn from a part of the labeled data (training) to predict the class label of the unseen data, called test data. The rationale of using the classifier in this study was to evaluate which subset of the selected genes by the two above mention feature selection techniques can lead to more accurate discrimination between three groups of NL, MCI, and AD. Since each classification technique’s performance might be different in terms of accuracy, we used three different classifiers, namely support vector machine (SVM), adaptive boosting (AdaBoost), and K-Nearest Neighbor (KNN).

SVM is a highly preferred and robust supervised machine learning algorithm that usually is used for solving binary classification problems. The SVM algorithm performs the classification by finding a hyperplane that maximizes the margin between two classes. The SVM performance depends on two primary hyperparameters named cost (C) and gamma (γ). The C is the penalty parameter, which indicates how much the algorithm cares about misclassification by adding a penalty for each misclassified sample, and gamma determines the extent of the effect of a single training example. In this study, we use an SVM with a linear kernel, and we defined the C=1 and the value from the equation of 1 / (n_features × X.var()) for the gamma (X denotes the input values).

Boosting algorithms have recently attracted significant attention because of their low-complexity implementation, good generalization, and processing speed [16]. These types of algorithms usually have higher accuracy than some of the conventional machine learning algorithms because of their ability to convert weak learners into strong learners. They are not only considered as a beneficial technique for solving binary classification tasks but also are useful in solving multi-class classification problems. A booster is composed of a set of training samples (*x*_1_, *y*_1_),… (*x*_n_,*y*_n_), in which all the outcomes (*y*n) are labeled associated with observations (*x*n) [17]. Adaptive boosting, which is also named Adaboost, is considered as a meta-estimator classifier that aims to estimate the Bayes classifier iteratively by incorporating many weak classifiers and inaccurate rules, which can finally turn a weak classifier into a robust and strong classifier. In this study, we determined a Decision Tree Classifier as a base estimator for the AdaBoost algorithm, and also, we defined the number of the estimators as 45.

The K-Nearest Neighbor (KNN) was the third classifier we used in this study. The KNN is considered the most acceptable choice when very little or no prior knowledge present about the data distribution [18]. The term K in KNN is a parameter that refers to how many numbers of nearest neighbors are chosen for the KNN algorithm. For each sample, the algorithm calculates the distance with respect to K and classifies it into the nearest distance neighbor class. In this work, we selected the K to be 5.

#### 5) Gene ontology (GO) enrichment analysis

The goal of all previous steps was to determine which subset of the genes is differentially expressed between three conditions of NL, MCI, and AD and which subset of the genes can lead to the most accurate discrimination between the three mentioned groups. However, the output of the analysis methods often leads to a long list of DEGs, which makes the interpretation a bit challenging. GO enrichment analysis can be a very useful approach to tackle this issue and to interpret and classify the DEGs’ function at the molecular and cellular levels. Gene Set Enrichment Analysis (GSEA) is a computational method developed for gene expression analysis from microarray data [19]. The aim of the GSEA is to look for the expression changes in predefined gene sets. In other words, GSEA uses a list of ranked genes to evaluate their distribution on predefined genes set, such as MSigDB [20], by determining an enrichment score (ES) for each set of genes [21]. In this study, we used GSEA software version 4.1.0 [19, 22] for further analysis. To do so, we selected c5.all.v7.2.symbols.gmt (Gene ontology) as the gene set database, the number of permutations was set on 1000, and we considered the dataset as is in the original format. Since the ADNI used the Affymetrix Human Genome U219 Array (Affymetrix (www.affymetrix.com), Santa Clara, CA) for expression profiling, we used HG_U219 as the chip platform. Further, we set the enrichment statistic on weighted, max size for excluding the larger sets on 500, and min size for excluding the smaller sets on 2.

## Results

Unpaired two-sided Student’s t-test (FDR adjusted p-value <0.05) was used to identify the DEGs from the original expression dataset. The t-test between the pair of NL vs. MCI resulted in 1559 genes, 5005 genes for the pair of NL vs. AD, and 5949 genes for MCI vs. AD. Since DEGs analysis between three different pairs was led to a long list of genes, to summarize the relationship between the genes and also to identify the commonly expressed genes between NL, MCI, and AD, we used the Venn diagram. Finally, as shown in fig.5, 39 genes were identified by the Venn diagram as the commonly expressed genes. It is worth mentioning that nine genes were removed from the commonly expressed gene list because of the repetition, and the 30 remaining genes were described in Table 2.

**Figure 5.**
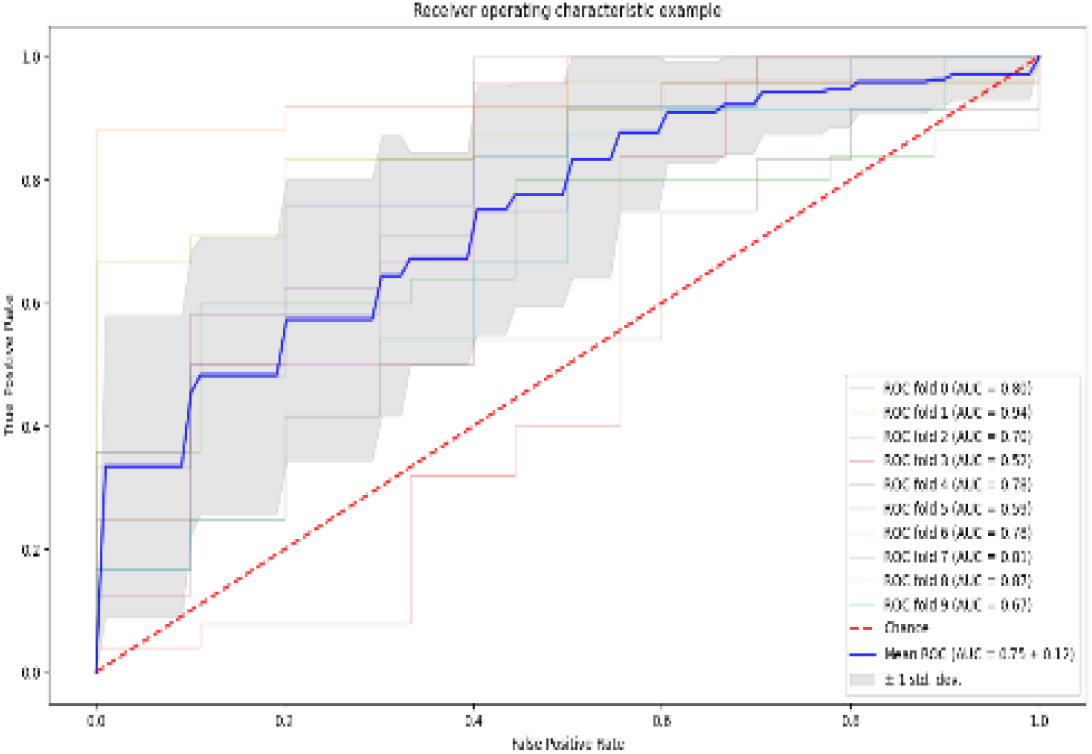
ROC curve for RFE and SVM

**Table 2.** List of DEGs

| ProbeSet | LocusLink | Symbol | P_value |
| --- | --- | --- | --- |
| 11746877_a_at | LOC83463 | MXD3 | 0.000667 |
| 11722971_a_at | LOC9586 | CREB5 | 0.001372 |
| 11733513_a_at | LOC79971 | WLS | 0.001372 |
| 11760084_x_at | LOC254896 | AC107959.4 | 0.001372 |
| 11740560_x_at | LOC8439 | NSMAF | 0.001455 |
| 11761745_at | - | IGSF6 | 0.002528 |
| 11752332_x_at | LOC4210 | MEFV | 0.003398 |
| 11729533_s_at | LOC161253 | REM2 | 0.003851 |
| 11721632_a_at | LOC27346 | TMEM97 | 0.003981 |
| 11738921_at | LOC58160 | NFE4 | 0.004019 |
| 11747038_a_at | LOC254013 | METTL20 | 0.004019 |
| 11756516_a_at | LOC23761 | PISD | 0.005359 |
| 11747006_a_at | LOC387755 | INSC | 0.006366 |
| 11730579_a_at | LOC57520 | HECW2 | 0.006686 |
| 11739880_a_at | LOC169792 | GLIS3 | 0.006719 |
| 11738864_a_at | LOC145447 | ABHD12B | 0.007282 |
| 11750058_a_at | LOC55284 | UBE2W | 0.007282 |
| 11745206_x_at | LOC5887 | RAD23B | 0.007721 |
| 11723545_a_at | LOC5337 | PLD1 | 0.008008 |
| 11737032_x_at | LOC222235 | FBXL13 | 0.008474 |
| 11740041_at | LOC57194 | ATP10A | 0.008474 |
| 11728950_at | LOC132228 | LSMEM2 | 0.010634 |
| 11759103_at | - | CYP4F3 | 0.010818 |
| 11758554_s_at | LOC64326 | RFWD2 | 0.01092 |
| 11751762_s_at | LOC26512 | INTS6 | 0.011011 |
| 11751370_a_at | LOC51762 | RAB8B | 0.011077 |
| 11751989_a_at | LOC3696 | ITGB8 | 0.011359 |
| 11716818_a_at | LOC11326 | VSIG4 | 0.011663 |
| 11738159_x_at | LOC125965 | COX6B2 | 0.011663 |
| 11747940_a_at | LOC127435 | PODN | 0.012904 |

### B. Most important selected genes

The top 10 genes were selected by using the two well-known feature selection methods (RFE and LassoCV). At the first step, all the 30 genes from the intersection of NL, MCI, and AD were ranked based on their coefficient and importance using RFE. The V-set immunoglobulin-domain-containing 4 (VSIG4) ranked first as the most important gene that can be used for classification between CN, MCI, and AD. WLS (Wnt Ligand Secretion Mediator), TMEM97 (Transmembrane Protein 97), PLD1 (Phospholipase D1), and COX6B2(Cytochrome C Oxidase Subunit 6B2) we the next four important genes which were selected by RFE

Moreover, GLIS3 (GLIS Family Zinc Finger 3), ETFBKMT (Electron Transfer Flavoprotein Subunit Beta Lysine Methyltransferase), MXD3 (MAX Dimerization Protein 3), PODN (Podocan), and ITGB8 (Integrin Subunit Beta 8) genes were the next five important genes were selected by RFE, respectively.

In the same way, the LassoCV considered the ITGB8 as the most prominent gene, while this gene was ranked 10th by RFE. GLIS3, TMEM97, METTL20, PODN, and MXD3 were the other genes determined as ranked second to fifth and seventh, respectively. These five mentioned genes were also selected by RFE as the important genes. However, four other genes were selected by LassoCV, which did not consider as high-ranked genes by RFE. ATP10A (ATPase Phospholipid Transporting 10A, INTS6 (Integrator Complex Subunit 6), RAB8B (RAB8B, Member RAS Oncogene Family), and NFE4 (Nuclear Factor, Erythroid 4) were the genes ranked sixth, and eight to tenth respectively by LassoCV. The results of the two feature selection methods are shown in Table 3.

**Table 3.** Top 10 genes selected by two feature selection methods

| Rank | Genes Selected by Feature Selection Techniques |  |
| --- | --- | --- |
|  | RFE | LassoCV |
| 1 | V-set immunoglobulin-domain-containing 4 (VSIG4) | Integrin Subunit Beta 8 (ITGB8) |
| 2 | Wnt Ligand Secretion Mediator (WLS) | GLIS Family Zinc Finger 3 (GLIS3) |
| 3 | Transmembrane Protein 97 (TMEM97) | Transmembrane Protein 97 (TMEM97) |
| 4 | Phospholipase D1 (PLD1) | Mitochondrial Lysine Methyltransferase (METTL20) |
| 5 | Cytochrome C Oxidase Subunit 6B2 (COX6B2) | Podocan (PODN) |
| 6 | GLIS Family Zinc Finger 3 (GLIS3) | ATPase Phospholipid Transporting 10A (ATP10A) |
| 7 | Electron Transfer Flavoprotein Subunit Beta Lysine Methyltransferase (ETFBKMT) | MAX Dimerization Protein 3 (MXD3) |
| 8 | MAX Dimerization Protein 3 (MXD3) | Integrator Complex Subunit 6 (INTS6) |
| 9 | Podocan (PODN) | Ras-related protein Rab-8B (RAB8B) |
| 10 | Integrin Subunit Beta 8 (ITGB8) | Nuclear Factor, Erythroid 4 (NFE4) |

As mentioned above, the five genes of ITGB8, GLIS3, TMEM97, PODN, and MXD3 were the genes that both feature selection techniques considered as the high importance features for classification between NL, MCI, and AD.

After the feature selection step, since the RFE and LassoCV selected different subsets as the most important genes, we used three binary classification methods (SVM, AdaBoost, and KNN) with 5-fold cross-validation to evaluate the efficiency of the top 10 selected genes by LassoCV and RFE.

As can be seen in Table 4, the best classification result (mean AUC of 0.77 ± 0.08) was achieved for the pair of NL vs. AD when the features selected by LassoCV were fed to the SVM classifier. The SVM also resulted in a relatively close mean of AUC (0.75 ± 0.12) for classification between NL and AD using the RFE selected genes. The ROC curve (receiver operating characteristic curve) is a graph showing the performance of a classification model at all classification thresholds. The ROC of both results is shown in figures 6 & 7. Although the highest result for classification between NL and AD was achieved by the genes were selected using LassoCV, based on the results, it seems the selected genes using RFE were led to better classification results for NL vs. MCI with the mean AUC of 0.64 ± 0.04, and the mean AUC of 0.62 ± 0.08 for MCI vs. AD.

**Table 4.** Classification results for all of the states compared two by two applying SVM, AdaBoost, and KNN

| Compared Groups | Classifier | Mean AUC ± std | |
| --- | --- | --- | --- |
|  |  | LassoCV | RFE |
| NL vs. MCI | SVM | $0.59 \pm 0.05$ | $0.64 \pm 0.04$ |
| | AdaBoost | $0.55 \pm 0.05$ | $0.57 \pm 0.04$ |
| | KNN | $0.56 \pm 0.07$ | $0.55 \pm 0.05$ |
| NL vs. AD | SVM | $0.77 \pm 0.08$ | $0.75 \pm 0.12$ |
| | AdaBoost | $0.63 \pm 0.12$ | $0.62 \pm 0.14$ |
| | KNN | $0.64 \pm 0.11$ | $0.60 \pm 0.09$ |
| MCI vs. AD | SVM | $0.57 \pm 0.13$ | $0.56 \pm 0.09$ |
| | AdaBoost | $0.58 \pm 0.11$ | $0.62 \pm 0.08$ |
| | KNN | $0.56 \pm 0.08$ | $0.59 \pm 0.09$ |

**Figure 6.**
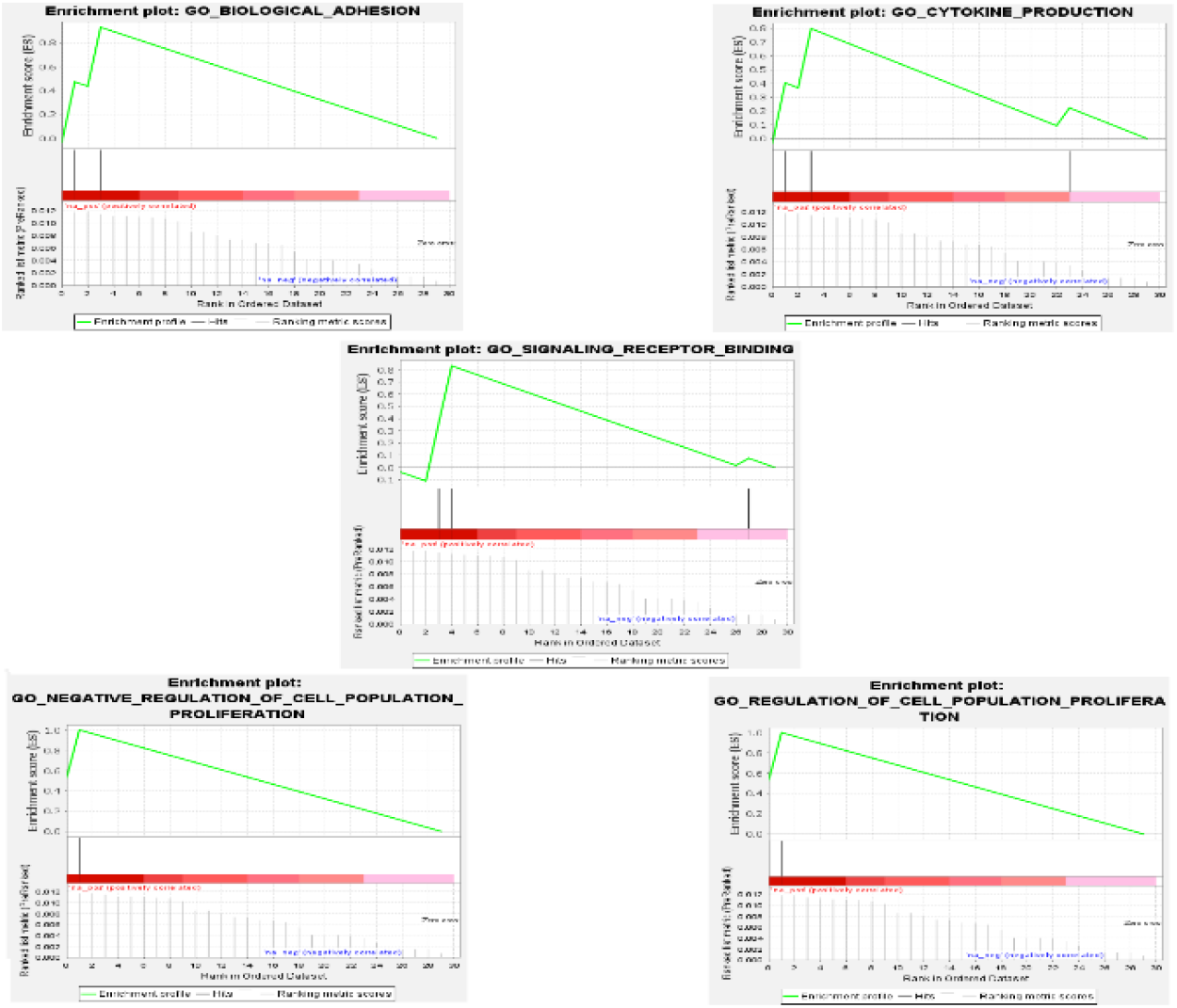
The enrichment plot of all the enriched GO terms.

We also considered the six genes that were common between RFE and LassoCV for further analysis to evaluate their efficiency for classification between NL, MCI, and AD. The mean AUC of 0.75 ± 0.12 was achieved for classification between NL and AD by SVM, which was similar to the result of the classification between NL and AD using the RFE selected genes. The other classification results (NL vs. MCI and MCI vs. AD) were not significant enough to mention.

### C. Gene ontology (GO) enrichment analysis

Thirty intersected differentially expressed genes between NL vs. AD, NL vs. MCI, and MCI vs. AD that were ranked according to their p-values used for the GO enrichment analysis. The GO enrichment test was performed and GO terms with the p-values <0.05 were considered as the significant enriched GO terms. It is worth mentioning that by considering the q-value <0.05, none of the GO terms was significantly enriched. Nevertheless, by considering the p-value <0.05, representative GO terms enriched via four genes, namely PODN, VSIG4, ITGB8, RAB8B, were as follows: regulation of cell proliferation (core enrichment: PODN, VSIG4), negative regulation of cell population proliferation (core enrichment: PODN, VSIG4), signaling receptor binding (core enrichment: ITGB8, RAB8B), biological adhesion (core enrichment: VSIG4, ITGB8), and cytokine production (core enrichment: VSIG4, ITGB8). The enrichment plot of all the enriched GO terms is illustrated in Figure 8.

## Discussion

The main goal of this study was to find the most significant and relevant genes that can lead to the most accurate classification between normal, MCI, and AD patients, from the ADNI blood gene expression dataset. This research started with the ADNI data normality check. The t-test and Fold Change (FC) were used to identify DE genes for all three groups. The two-sided unpaired Student’s t-test with the significance level of 0.05 was implemented to check each two sample groups. Similar to other studies on the ADNI dataset, gene expression differences between samples in different classes were low. We selected DEGs based on adjusted p-values < 0.05 (FDR), and 1559 genes passed after comparing NL vs. MCI groups. Additionally, from the comparison between NL vs. AD and MCI vs. AD, 5005 and 5949 genes passed, respectively.

Then 39 common genes between all the pairs were selected using a Venn diagram. Nine duplicated genes were removed from further analysis, and the number of the final common genes was reduced to 30 genes. RFE and LassoCV methods selected the top 10 genes from the 30 genes, and they were used as input to the SVM, AdaBoost, and KNN binary classification methods. The best result was obtained from the comparison of NL vs. AD using the SVM classification with the mean AUC of 0.77 ± 0.08 for the LassoCV technique and 0.75 ± 0.12 for the RFE analysis method. Our proposed feature selection method, together with SVM classification, significantly outperformed T. Leel & Hyunju Lee’s results [4].

We perceived different enriched GO terms, and also, most of the selected genes directly or indirectly are related to AD by affecting on human nervous system.

In humans, the ITGB8 gene is a member of the integrin beta chain family that encodes integrin beta-8. Apart from cell-extracellular matrix (ECM) and cell-cell adhesion mediation [23], most β subunits contain an NPxY sequence in their cytosolic region which can interact with the cytoskeletal and signaling proteins containing a phosphotyrosine binding (PTB) domain [24]. Interestingly, these heterodimeric transmembrane receptors have been suggested to play a role in intracellular Ca2+ and protein kinases signaling and reorganization of cytoskeletal filaments. It has been shown that integrin and ECM are altered in physiological and pathological conditions such as memory, tumor, Alzheimer’s disease, and epilepsy [25, 26]. There are consistent reports of abnormal calcium signaling in Alzheimer’s disease [27]. Moreover, dysregulation of protein kinase signaling cascade promotes the development of AD and contributes to neuronal death [28]. These findings may suggest that dysregulation of ITGB8 expression could be related to these alterations, which leads to the development of Alzheimer’s disease.

The next important selected gene by our model was GLIS3 (GLI-similar 3) which is a member of the GLI-similar zinc finger protein family encoding for a nuclear protein. This protein has a critical role in both repression and activation of transcription. Interestingly, Genome-Wide Association Studies (GWAS) have shown that GLIS3 is a risk gene for Alzheimer’s disease endophenotype [29], and common variants within GLIS3 itself are associated with cerebrospinal fluid Tau [30], a biomarker in Alzheimer’s disease.

As a matter of fact, memory loss is one of the main symptoms of the AD and TMEM97 (transmembrane protein 97) as a gene that codes for the Sigma-2 receptor [31] is involved in calcium homeostasis [32, 33], which is critical for the excitatory imbalance that regulates learning and memory. In human post-mortem brain samples, TMEM97 has been presented in a higher proportion of synapses and close enough to amyloid-beta to be recognized as a potential synaptic binding partner [34].

Another top selected gene that seems to play a critical role in AD prediction is PODN (Podocan). PODN is a protein-coding gene that codes for a novel member of the small leucine-rich repeat protein family. It was shown that ECM molecules are regulators of essential cell functions such as migration and proliferation [35, 36], and the small leucine-rich repeat proteins found in the ECM are also effective modulators of cell phenotype [37]. It has been demonstrated that overexpression of podocan in Chinese Hamster Ovary (CHO) cells leads to a marked suppression of cell migration. Besides, cells transfected with podocan showed a modest decrease in Rho A activity compared to vector-transfected cells [38]. It is evident that Rho proteins play a crucial role in organelle development, cytoskeletal dynamics, cell movement, and other common cellular functions [39-41], and aberrant Rho-GTPase signaling leads to widespread neuronal network dysfunction and has been proposed to cause certain diseases, including AD. Recent immunohistological studies suggest that the subcellular localization of RhoA may be altered in AD brains [42].

The final gene selected as an important gene by both feature selection techniques was MXD3 (MAX dimerization protein 3), a protein in humans encoded by the MXD3 gene and contributes to normal neural development and brain cancer development [43, 44]. Several studies suggest that this gene plays a crucial role in metabolic rewiring and context-specific tumor suppression [45, 46]. Interestingly the MXD3 protein binds E-box sequences and increases cell proliferation at moderate MXD3 levels, but it can cause marked cell death and apoptosis at higher expression levels in medulloblastoma cells [44]; however, the role of the MXD3 gene in the development of AD is not well understood and needs more investigation.

## Conclusion

In this study, we investigated straightforward statistical and advanced feature selection-based analysis on human blood gene expression data to identify the differentially expressed genes that are mostly related to MCI and AD. Three classifiers were employed to differentiate between three groups of NL, MCI, and AD using the selected genes. In addition, GO enrichment analysis was performed to interpret and classify the selected genes’ function at the molecular and cellular levels. The analysis resulted in selecting the top 10 genes, which can lead to discrimination between two groups of NL and AD with an average AUC of 0.77 ± 0.08. Furthermore, the five genes selected by both feature selection techniques as the most important genes were discussed in detail. Moreover, gene ontology (GO) analysis showed that four genes were enriched with the GO terms of regulation of cell proliferation, negative regulation of cell population proliferation, signaling receptor binding, biological adhesion, and cytokine production. Although, at first glance, some of the selected genes may not seem to be directly related to AD or MCI, they can open a new window for further investigation, and they could be potential risk-factor genes for MCI and AD.

## Data Availability

The data we used is available online and can be download from the following link: “http://adni.loni.usc.edu/data-samples/data-types/genetic-data/ webiste.

